# A new multi-species Protein A-ELISA assay for plague diagnosis in humans and other mammal hosts

**DOI:** 10.1101/2021.09.21.21263536

**Authors:** Matheus Filgueira Bezerra, Camila Cavalcanti Xavier, Alzira Maria Paiva de Almeida, Christian Robson de Souza Reis

## Abstract

**Background:** The Hemagglutination assay (HA) is widely used in plague diagnosis, however, it has a subjective interpretation and demands high amounts of antigen and other immunobiological supplies. Conventional IgG-ELISA is limited by the need of specific conjugates for multiple plague hosts.

**Methods:** Thus, we developed an ELISA Protein A-peroxidase method to detect anti-F1 antibodies across several species, including humans. To determine the cut-off and performance rates, HA results from 288 samples (81 rabbits, 64 humans, 66 rodents and 77 dogs) were used as reference.

**Results:** Optimal conditions were found with 250ng/well of F1 and 1:500 serum dilution. Protein A-ELISA showed high repeatability and reproducibility. The positive/negative OD ratios were higher in Protein A-ELISA and there was no significant cross-reaction with other pathogenic yersiniae. The overall sensitivity/specificity, area under the curve and Kappa rates for Protein A-ELISA were 93.9/98.9%; 0.993 and 0.938, respectively. Similar results were observed in each species separately. There was a strong agreement between Protein A and IgG assays (kappa=0.973) in independent analysis (n=487).

**Conclusions:** Altogether, the Protein A-ELISA showed high performance when compared both to HA and IgG-ELISA, with a polyvalent single protocol that requires reduced amounts of antigen and can be employed to any plague hosts.

## INTRODUCTION

Plague is a flea-transmitted disease caused by the gram-negative bacterium *Yersinia pestis* and was responsible for at least three pandemics in the past [1]. Although nowadays plague can be treated with antibiotics, there is a lack of vaccines able to provide long-term immunity and this disease still threaten individuals living in remote places, close to wildlife hosts but distant from specialized healthcare services [2]. Human cases and deaths are recorded annually in several countries across Africa, Asia and the Americas [3]. Despite the declining incidence worldwide, the interest in plague is constant because of its potential to establish new epidemics and application as a biological weapon [2,4].

Although the rodents are the main plague reservoir, practically any mammal can be infected by *Y. pestis* and may take part in the dynamics of the infection [5]. An interesting feature of plague is that, under certain conditions, the disease is able to remain quiescent in the natural foci for decades and eventually reemerge among the wild fauna and spillover to human populations [6,7,8]. Due to this unique feature, it is of utmost importance to perform continuous monitoring of plague areas. In this regard, serological methods are an important surveillance tool, as it identifies not only animals with the active form of the disease, but also those previously exposed [9]. Most serological tests for plague are based on the detection of antibodies against the F1 capsular antigen, which is exclusive to *Y. pestis*, and highly immunogenic for humans and other mammals [10,11].

Given its polyvalence for sera from all taxonomic family groups, hemagglutination (HA) has been widely used for plague serological diagnosis for several decades. However, some commonly observed problems in HA, such as interpretations bias, cross-reaction with other infections, high consumption of F1 antigen and use of perishable biological supplies, led many laboratories to migrate to IgG ELISA tests [12-16]. On the other hand, conventional ELISA requires a specific anti-IgG conjugate and different optimization for each mammal species. Thus, there is a need of new diagnostic methods that can improve the diagnosis and epidemiological surveillance of human and animal plague across the globe [2,4,17].

Alternatively to immunoglobulin (anti-IgG) conjugates, the *Staphylococcus aureus* protein A has been proposed for diagnosis of other multi-host diseases due to its universal affinity for immunoglobulins from various species of domestic and wild mammals [18-20]. To tackle this gap, we proposed a Protein A-based indirect ELISA method, able to detect anti-F1 antibodies from humans and other plague hosts within a single protocol.

## METHODS

### F1 production

The F1 antigen was extracted from the A1122 *Y. pestis* strain, according to the protocol described by Chu [10], in a Biosafety level 3 facility. The purified product was then mixed with 2× Laemmli buffer (1:1) containing 5% β-mercaptoethanol, heated at 100 °C for five minutes and loaded onto a 20% polyacrylamide gel (Figure 1A). The F1 antigen was quantified using the NanoDrop One^C^ Microvolume UV-Vis Spectrophotometer (Thermo-Fischer, USA).

**Figure 1.**
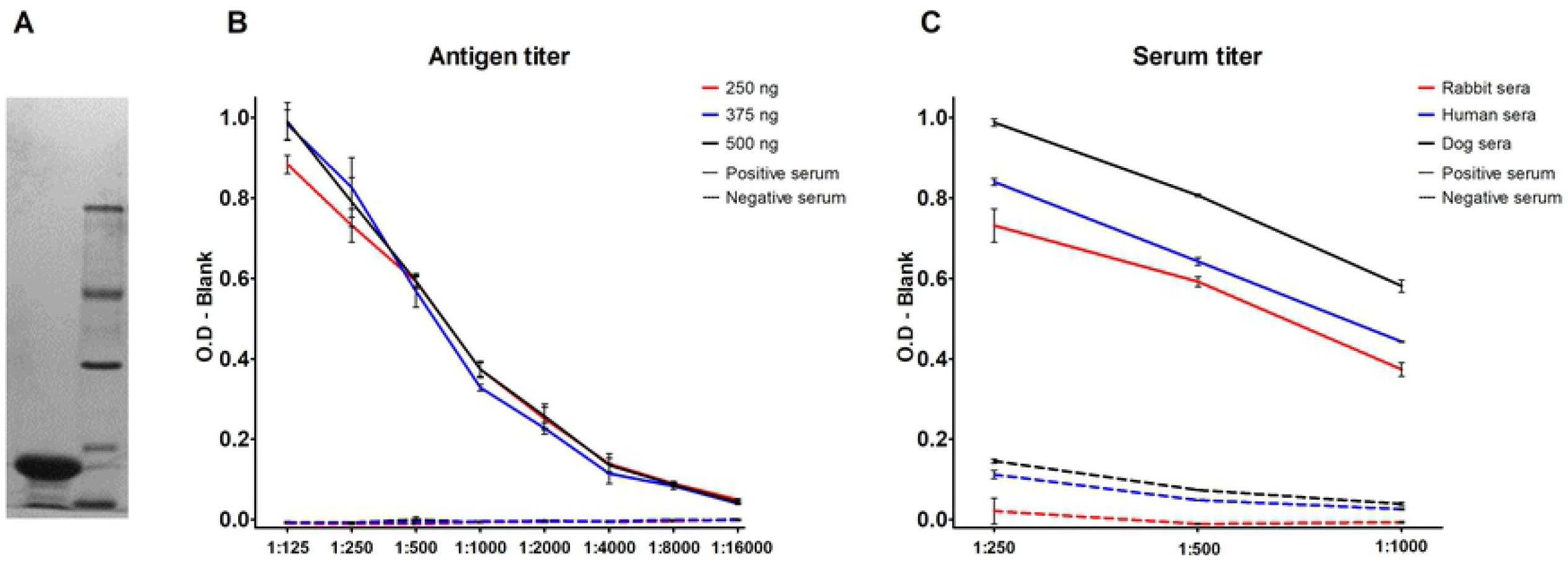
Standardization of Protein A-ELISA. F1 antigen purified from the A1122 *Y. pestis* strain in culture **(A)**. Three concentrations of F1 antigen (250 ng, 375 ng and 500 ng) were tested. The assay was optimized at the concentration of 250 ng of F1 antigen per well **(B)**. Graph with the optical densities from titrated sera in different species **(C)**.

**Figure 2.**
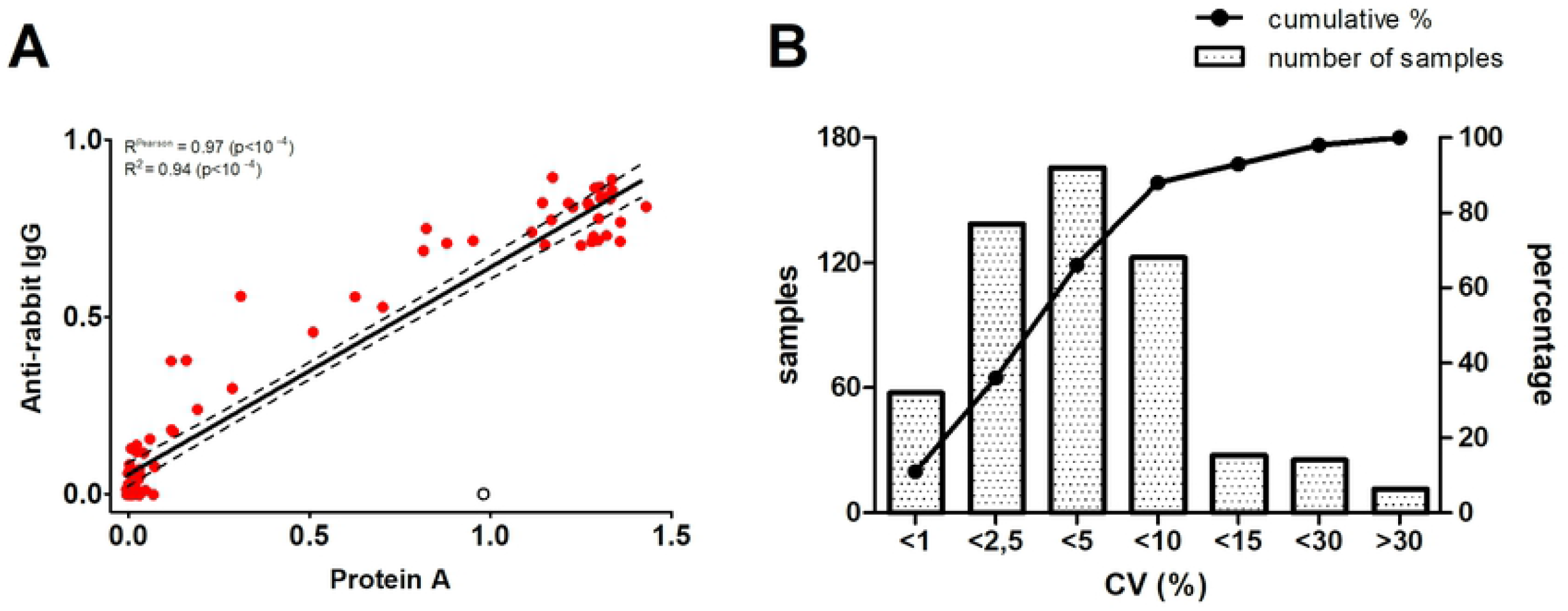
Linearity and precision of Protein A-ELISA. Good correlation (Pearson=0.97) was observed between the Protein A and IgG ELISA tests using control rabbit sera (n = 81). Dashed lines represent the 95% confidence interval and the white datapoint, an outlier samples excluded from correlation analysis **(A)**. From the total 553 samples tested for Protein A-ELISA, 88% had a coefficient of variation (CV) of the triplicates lower than 10% and 93% of the samples had a CV lower than 15%. The line shows the cumulative percentage of samples within the respective CV level and the bars show the absolute amounts of samples in each interval of CV value **(B)**.

### Human and animal samples

Initially, 288 sera (81 control rabbits, 64 humans, 66 wild rodents and 77 domestic dogs; total positives/negatives = 98/190) with well-characterized results for HA were used for cut-off determination and Protein A-ELISA validation. Next, 265 additional samples with unknown HA results nor used in cut-off determination were included to evaluate the agreement between the Protein A and IgG methods in ELISA. All sera were kindly provided by the Brazilian Plague Reference Service (SRP) and originated from the routine surveillance of the Brazilian plague areas including human cases, several rodent species and domestic carnivores (stray dogs) that pray on rodents [21,22].

### Rabbit immunization

Sera from rabbits immunized with formol-killed *Y. pestis* and other pathogenic Yersinia strains (whole-cells immunization) or with the purified F1 antigen, produced as previously described [23] for positive control in routine diagnosis and were kindly provided by the SRP. From the 37 positive control sera, 14 were from rabbits exposed to the reference EV76 or A1122 *Y. pestis* strains in independent experiments, 18 were from rabbits exposed to diverse Brazilian *Y. pestis* strains from the Fiocruz-CYP (http://cyp.fiocruz.br) bacterial cultures collection and five were from rabbits exposed to the purified F1 antigen (three native F1 and two recombinant F1, expressed in *E. coli*) [23]. Additionally, to evaluate whether the ELISA test would present cross-reaction with other Yersiniae, five rabbits immunized with distinct isolates of *Yersinia pseudotuberculosis* and two with *Yersinia enterocolitica* were included. The other 37 negative control rabbit sera were obtained from animals from the IAM facilities that did not underwent any experimental intervention.

### Ethics Statement

The production of immune sera for positive controls in rabbits is routinely produced following the local Animal Ethics Committees (CEUA/IAM) guidelines and supplies the laboratorial diagnosis of plague by the National Plague Reference Service (SRP–IAM). Sera from rodents, dogs and humans were obtained during the routine operations of the Brazilian plague surveillance program over the years and maintained in the serum samples collection of the Instituto Aggeu Magalhães.

### Protein A-ELISA and IgG-ELISA

The ELISA tests were adapted from previously established protocols [10,13]. Briefly, 96-well plates (Techno Plastic Products, Switzerland) were incubated overnight with 250 ng of F1 diluted in 100 µl of a 0,05 M, pH 9,6 carbonate-bicarbonate buffer per well. Next, the plates were washed twice with 500 µl of PBS per well (PW 40 Microplate Washer, Bio-Rad, USA) and blocked with 100 µl of a 10% solution of low-fat milk in PBS for one hour. After a double wash with 500 µl of PBS-T (Tween 20, 0.05%), 100 µl of serum samples diluted (1:500) in a 10% milk/PBS-T solution were incubated in the plate at room temperature for one hour and washed twice with 500 µl of PBS-T. A 100 µl of Protein A–Peroxidase from *Staphylococcus aureus*/horseradish (Sigma-Aldrich, USA) or goat anti-human, rabbit or dog-peroxidase (Kirkegaard & Perry Laboratories, USA) diluted in 10% milk/PBS-T solution (1:10.000 and 1:2.500, respectively) were added and incubated at room temperature for one hour and washed twice with 500 µl of PBS-T. Finally, 150 µl of 2 mg/mL OPD (o-phenylenediamine dihydrochloride; Sigma– Aldrich) and 1:10^3^ H2O2 diluted in citrate-phosphate buffer (pH=5.0) was incubated in each well for 30 minutes at room temperature in a dark environment. The reaction was stopped by the addition of 100 μl 2.5 M sulfuric acid (H2SO4) per well and plates were read at the optical density of 490nm (iMark(tm) Microplate Absorbance Reader, Bio-Rad, USA).

All samples were measured in triplicates and the background (blank) optical density (OD) from each plate was subtracted from the average sample OD. Distinct concentrations of F1 antigen, peroxidase conjugates and sera dilution were tested to determine optimal conditions. The cut-offs were determined according to the best specificity/sensitivity (Youden’s index) from the ROC curve. A distinct cut-off was calculated for each peroxidase conjugate. Since the rodent samples included rather heterogeneous range of wild species [21], we could not test them for IgG-ELISA.

### Hemagglutination assay (HA)

The hemagglutination (HA) assay was performed as described previously [10]. In short, the F1 antigen was immobilized onto sheep red blood cells (SRBC) previously fixed with glutaraldehyde and tannic acid. Next, the F1-coated SRBC (25 µL/well) were incubated with the test serum serially diluted in eight wells starting from 1/4 in HA (0.85% saline + normal rabbit serum) buffer. The specificity of HA was accessed by the hemagglutination inhibition (HI). The test is considered positive when the HA endpoint is depressed by three or more HI dilutions (titers > 1/16 are considered positive).

### Statistical Analysis

The HA test, which is routinely used in the SRP, was used as the gold standard to calculate Protein-A ELISA and IgG ELISA performance rates. Sensitivity, specificity, accuracy and confidence intervals were calculated using the https://www.medcalc.org platform. Receiver Operating Characteristic (ROC) curves, area under the curve (AUC), scatterplots and correlations were calculated with the GraphPad Prism version 5 software. Pearson test was used to measure the correlation between ODs from distinct tests and Mann-Whitney test was used to compare OD means. The intra and inter-assay variability was measured using the coefficient of variation (CV) from one serum from a rabbit immunized with the A1122 *Y. pestis* strain and one negative rabbit serum. Samples were tested in eight replicates within runs and across six experiments in non-consecutive days.

The Kappa test was initially applied to determine the agreement rate between the ELISA and HA tests (n=288) and then, between Protein A-ELISA and IgG-ELISA (n=487). The index was calculated using the Quickcalc Graphpad tool (https://www.graphpad.com/quickcalcs/kappa2). Statistical tests were applied with a 95% confidence interval.

## RESULTS

### Standardization of Protein A-ELISA and IgG-ELISA

The optimal conditions were determined for Protein A-ELISA by evaluating separately distinct amount of F1 antigen per well and serum dilutions. There was no significant difference between the ODs by using the amounts of 250, 375 and 500 ng per well (Figure 1B). Thus, we decided to establish the lowest amount (250 ng per well) for the subsequent experiments. By testing three serum dilutions (1: 250; 1: 500 and 1: 1000), the 1:500 dilution showed high ODs for positive samples and low background for negative samples (Figure 1C). For optimization of the three IgG-ELISA tests, we maintained the amount of F1 antigen (250 ng/well) and sera dilution (1:500) previously established for Protein A-ELISA and tested four dilutions for IgG conjugate (1:1250; 1:2500; 1:5000 and 1:10000). We found the best positive/negative ratios at the 1:2.500 dilutions for all IgG conjugates (Supplementary Figure 1).

### Comparing ODs, cut-offs and cross-reaction between Protein A and IgG ELISAs

Whilst a single cut-off was established for Protein A-ELISA considering the best Youden’s index possible across all tested species, individual cut-offs were established for IgG anti-rabbit, anti-human and anti-dog ELISAs (Table 1 and Figure 3 A-B). We observed low background signals in negative samples for Protein A, anti-rabbit and anti-human IgG conjugates, but a rather marked background in anti-dog IgG conjugate, resulting in a narrower window of opportunity for cut-off.

**Figure 3.**
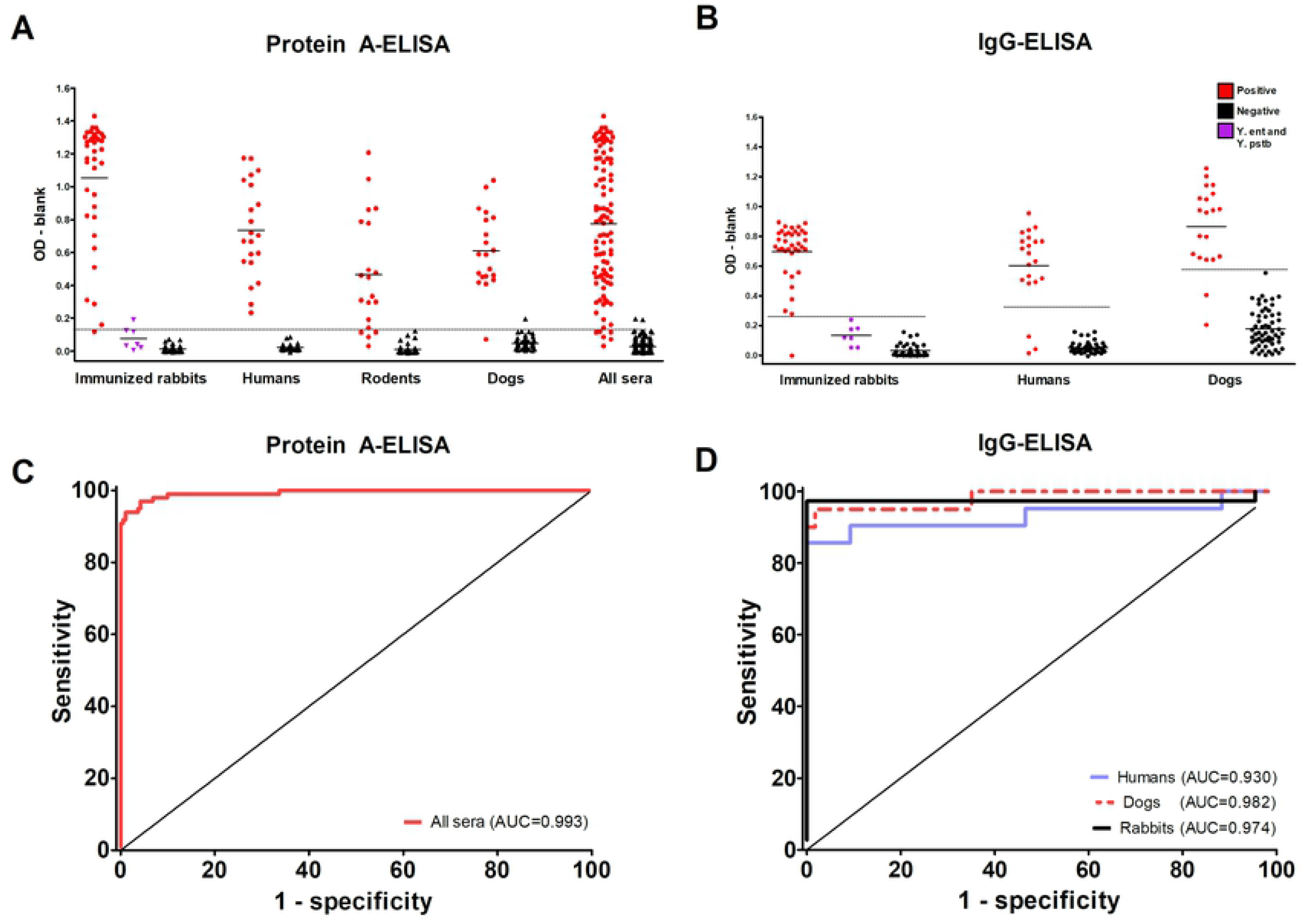
Validation of the diagnosis of plague by Protein A-ELISA and IgG-ELISA in HA-tested sera. Comparison between ODs from positive and negative sera for Protein A-ELISA. The cut-off in Protein A-ELISA was 0.130 for all species **(A)**. For IgG-ELISA, **t**he cut-offs were 0.258 (rabbits), 0.320 (humans) and 0.573 (dogs). All sera were previously tested for hemagglutination **(B)**. Area under the curve (AUC) for protein A **(C)** and IgG **(D)** conjugates. AUC values close to 1.0 indicate good test performance.

Overall, the average ODs from positive samples were significantly higher than the ODs from negative samples both in protein A and in IgG tests (Mann-Whitney test p < 10^−3^). The ratios between the OD means from positive and negative samples were considerably higher for the protein A conjugate (all samples = 28.7; rabbit = 81.1; human = 34.2; rodent = 51.7 and dog = 12.8), when compared to anti-rabbit (22.4), anti-human (11.8) and anti-dog (4.9) IgG conjugates. OD ratios and averages are shown in Table 1 and Supplementary Table 1, respectively.

We observed a good degree of correlation between ODs from Protein A and IgG-ELISA methods (Figure 2A). The mean ODs across six assay runs in different days were 1,392 (±0,069) for the positive rabbit serum (A1122) and 0,046 (±0,005) for the negative control, with a coefficient of variation (CV) of 4.9% and 10.8%, respectively. The repeatability, determined by eight intra-assay replicates, showed a CV of 0.5% for the positive anti-*Y. pestis* A1122 serum and 4.6% for the negative serum. Moreover, the intra-assay analysis of the triplicates from all samples tested for Protein A-ELISA revealed that 93% of the samples had a CV lower than 15% (Figure 2B). Of interest, only 2% of the samples had an CV above 30% and those were negative samples with ODs close to the lower detection limit, where small numeric variations imply in high CVs (Supplementary Figure 2).

To evaluate whether these ELISA methods would present cross-reaction with other pathogenic yersiniae, sera from seven rabbits previously immunized with *Y. pseudotuberculosis* (five) or *Y. enterocolitica* (two) strains were tested. Although the average ODs from these sera were slightly higher than other negative samples (protein A: 0.076 *versus* 0.014 and IgG: 0.133 *versus* 0.031), only one from the seven tested samples (*Y. enterocolitica*) presented a false-positive result for Protein A-ELISA (Figure 3 A-B).

### Performance of Protein A-ELISA and IgG-ELISA

Taking into consideration the HA results for 98 positive and 190 negative reference samples, the analysis of protein A and IgG ELISAs performance rates revealed high sensitivity, specificity and accuracy rates for both methods (Table 1). The Protein A-ELISA test had two false-positives (one rabbit and one dog) and six false negatives (four rodents, one rabbit and one dog), with an overall sensitivity of 93.9% and specificity of 98.9%. On the other hand, the IgG-ELISA test had six false-positives (three humans, two dogs and one rabbit), with a sensitivity of 97.4% and specificity of 100% for rabbits, 93% and 100% for humans and finally, 90% and 100% for dogs. Confirming these findings, the ROC curves from both protein A and IgG methods showed high area under the curve (AUC) rates. Whereas the overall and species-specific AUCs from protein A-ELISA tests remained above 0.990, AUCs from IgG-ELISA ranged from 0.930 to 0.982 (Figure 3 C-D, Supplementary Figure 3).

The Kappa test was initially applied to measure the degree of reliability between the ELISA tests and the HA (n=288 for protein A and n=222 for IgG). Excellent agreement rates were observed in samples from all species (Table 1). Next, we included new 265 independent samples with unknown HA results (and not used in cut-offs calculation) and calculated the kappa index to measure the agreement between Protein A and IgG ELISAs (Table 2). From the 487 samples, 84 were positive in both tests, 398 were negative in both tests, eight were positive for protein A but negative for IgG and five were positive for IgG but negative for protein A. Kappa coefficient for all species was 0.905 (0.854-0.956), for rabbits: 0.925 (0.842-1.000), for humans: 0.914 (0.818-and for dogs: 0.850 (0.741-0.959). The p-values were < 0.05 in all tests.

## DISCUSSION

The gold standard for plague diagnosis is the identification and isolation of the *Y. pestis* in bacteriological cultures from the clinical specimen. However, as proper diagnosis is often not feasible due to the acute progression of the disease and geographic isolation of cases, patients frequently receive treatment without laboratory results [2-4]. Therefore, serological testing is of most importance for plague diagnosis and surveillance activities, as it can detect not only active infections in humans and other hosts, but also retrospectively identify individuals exposed to the bacteria [9,10]. In this scenario, serological surveillance must consider a wide variety of mammals to be tested, such as rodents and other small mammals, domestic (dogs and cats) and wild carnivores that pray on rodents [5,12,21,22,24].

Here, we describe a Protein A-based approach designed to overcome some limitations faced by routine laboratories when using other serological methods, such as HA (subjective interpretation, high consumption of antigen, perishable reagents) and conventional ELISA (requires specific IgG-peroxidase conjugate, cut-off calculation and positive controls for each species). While the protocol here established requires 750 ng of F1 antigen per tested sample (using triplicates), HA spends 20,000 ng of F1 per tested sample (considering the standard eight dilutions according to Chu [10], resulting in the use of approximately twenty-seven times more antigen per sample. This difference can be particularly relevant for plague diagnosis given the complexity and costs of producing and purifying F1 from extensive *Y. pestis* culturing in biosafety level 3 (BSL3) laboratories [23].

Throughout a broad range of host species hereby tested, the Protein A-ELISA method showed high sensitivity, specificity and reproducibility rates even with a single cut-off value for all species. Corroborating these findings, the analysis of the ROC curve showed AUCs above 0.990 in all groups tested in protein A-ELISA, while AUCs from IgG-ELISA ranged from 0.930 to 0.982. The Cohen’s Kappa test revealed high agreement rates for this protocol when compared to HA (n=288) and IgG-ELISA (n=487). Since human cases of plague have not been reported in Brazil since 2005, we were not able to estimate positive/negative predictive values [25].

Remarkably, we observed a good correlation between ODs from Protein A and anti-IgG, with higher positive/negative OD ratios in the Protein A-ELISA test, which allows a safer window of opportunity for cut-off determination between positive and negative samples. Of note, little cross-reaction was observed in sera from rabbits immunized with other pathogenic yersiniae. Interestingly, whilst negative samples showed low background signals in Protein A, anti-rabbit and anti-humans IgG conjugates, a rather marked background in anti-dog IgG conjugate was observed. This could be associated to the non-specific agglutination routinely observed in sera from dogs in diagnosis by HA.

Although remaining detectable in humans for several years after infection, antibodies against *Y. pestis* can only be detected from the fifth day of infection by HA and from the eighth day by IgG-ELISA [15,26,27]. Previous studies demonstrate that in addition to its universal affinity for immunoglobulins (IgG) from almost all mammals, protein A can also bind to IgA and IgM and possibly, detect infections in earlier stages of infections, before serum conversion to IgG [19]. However, this hypothesis is yet to be tested in the context of serodiagnosis of plague.

Altogether, we validated a new indirect ELISA test that is sensitive, specific and reproducible, with a single protocol that can be used for both diagnosis of plague in humans and epidemiological surveillance in animal reservoirs from active foci.

## Data Availability

Further details on the dataset can be provided by authors upon request.

## ACKNOWLEDGMENTS

We are thankful to the Brazilian National Plague Reference Service staff for providing the serum samples to this study.

---

**Supplementary Figure 1.**
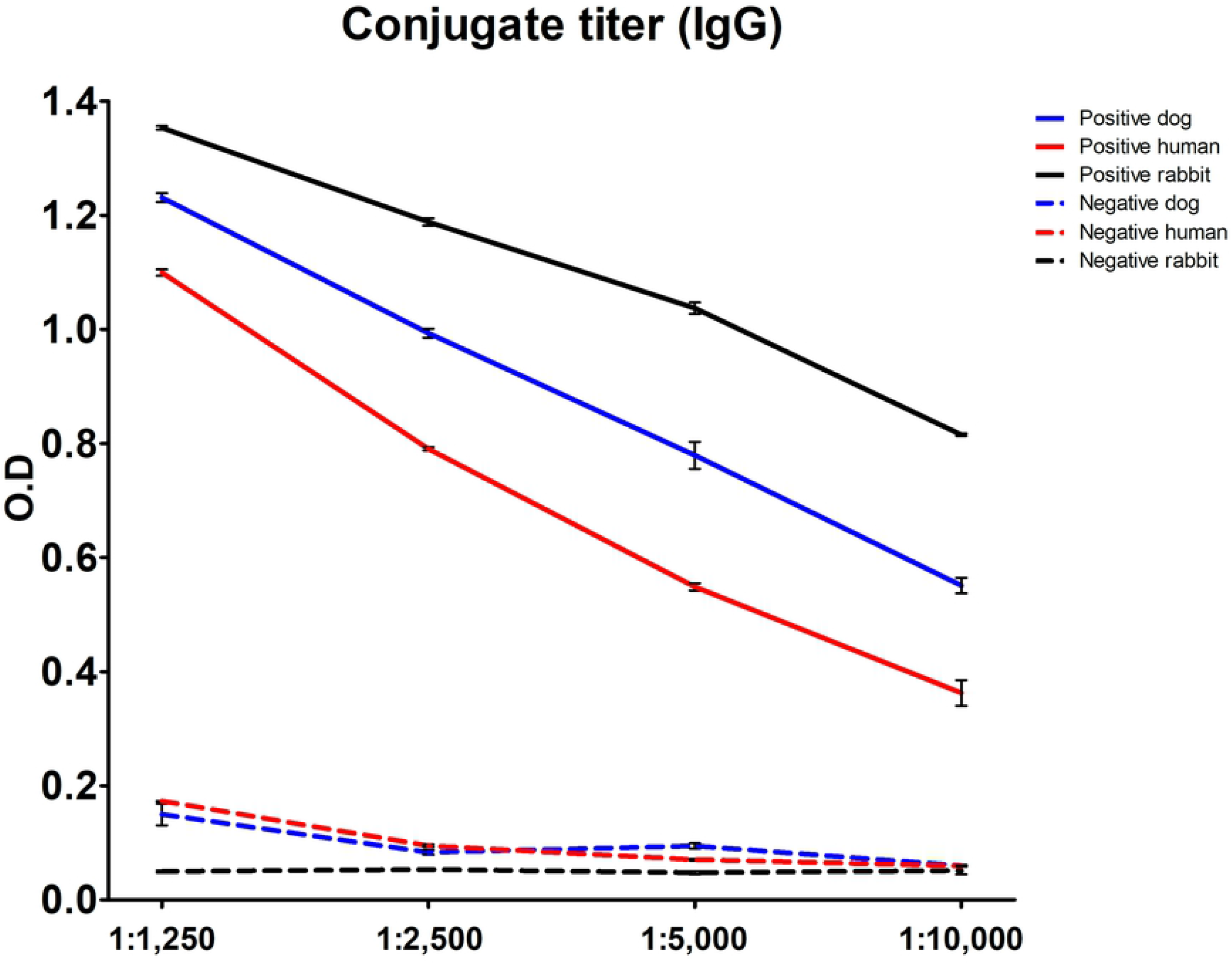
Standardization of IgG-ELISA. Four distinct titers were tested for each anti-IgG conjugates in triplicate for one positive and one negative serum from each species. Antigen concentration and sample titer were the same from the previously established in the Protein A-ELISA assay.

**Supplementary Figure 2.**
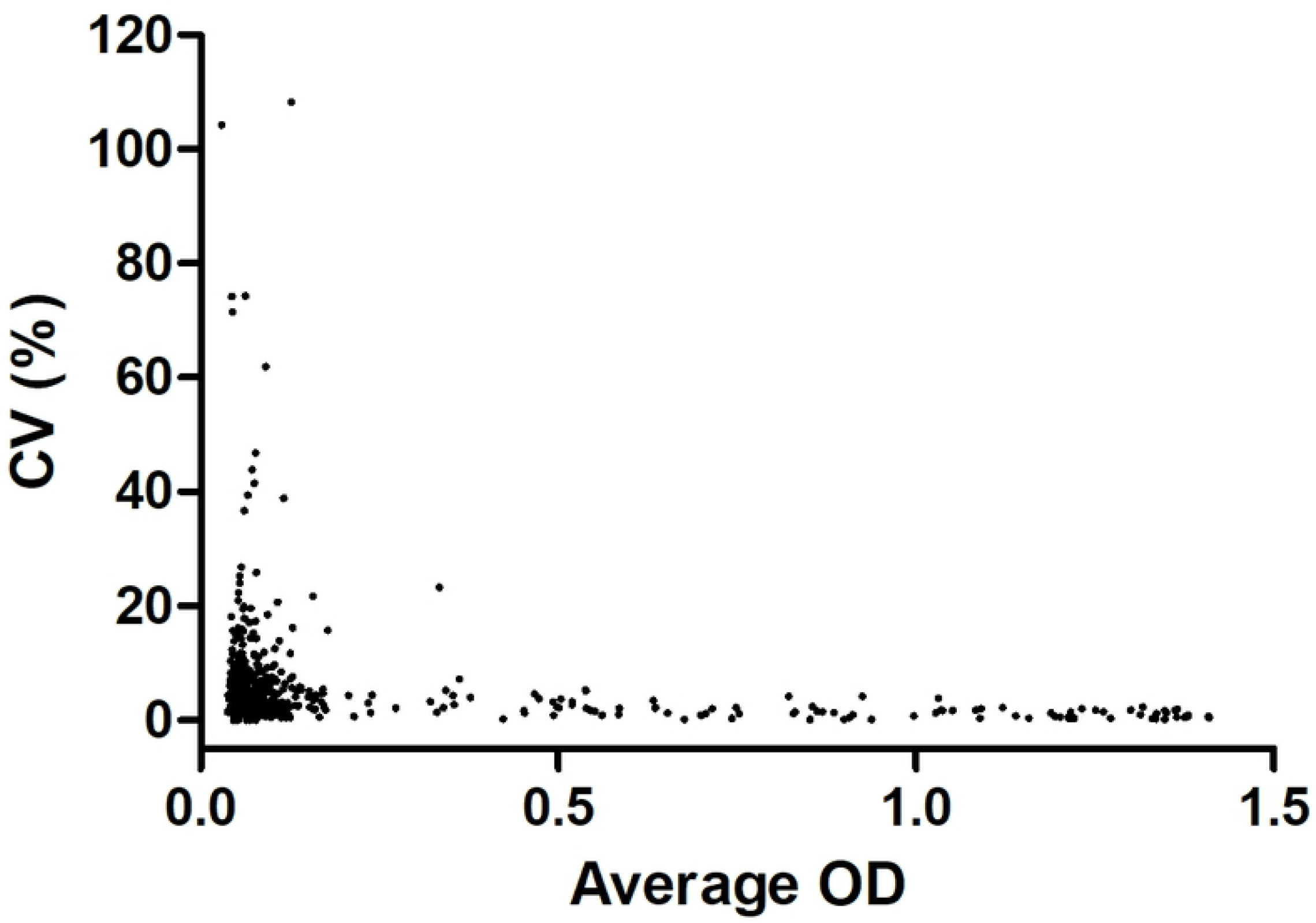
Coefficient of variation in Protein A-ELISA according to the average OD. The majority of the samples tested for Protein A-ELISA (n=553) had triplicates with low CVs. Higher variation was found in negative samples, with the ODs close to the lower detection limit, where small numeric variations implies in high CVs.

**Supplementary Figure 3.**
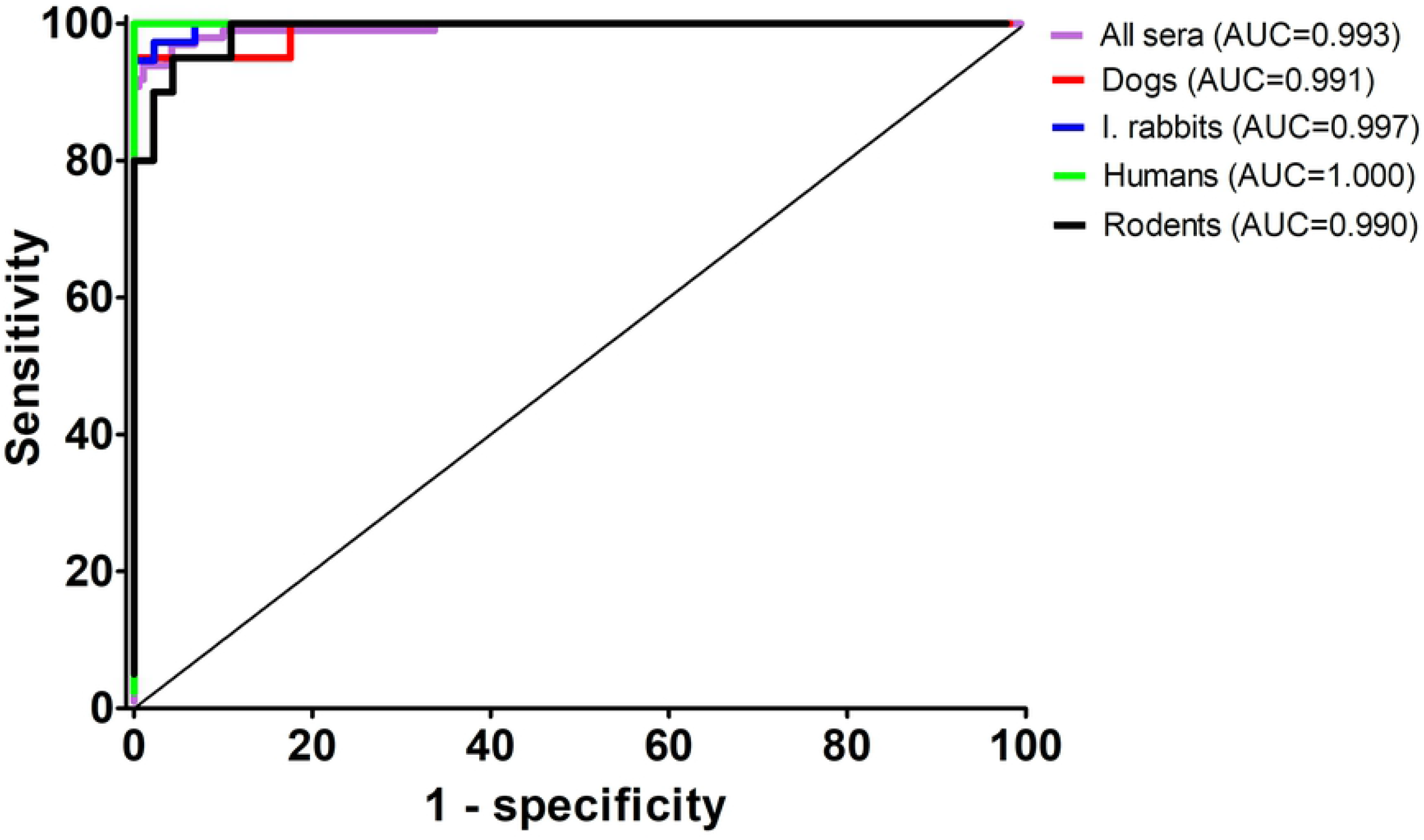
ROC curves for Protein A-ELISA. Stratified ROC curves and area under the curve (AUC) for protein A-ELISA according to each evaluated species.

